# Comparative immunogenicity of heterologous versus homologous 3rd SARS-CoV-2 vaccine doses in kidney transplant recipients

**DOI:** 10.1101/2022.01.25.22269778

**Authors:** Tina Thomson, Maria Prendecki, Sarah Gleeson, Paul Martin, Katrina Spensley, Charlotte Seneschall, Jaslyn Gan, Candice L. Clarke, Shanice Lewis, Graham Pickard, David Thomas, Stephen P. McAdoo, Liz Lightstone, Alison Cox, Peter Kelleher, Michelle Willicombe

## Abstract

**Background:** Solid organ transplant recipients have attenuated immune responses to SARS-CoV-2 vaccines. Emerging evidence suggests at least equivalent immunogenicity of heterologous compared with homologous vaccine regimens in the general population. In this study, we report on immune responses to 3^rd^ dose BNT162b2 vaccines in transplant recipients either primed with ChAdOx1 or BNT162b2.

**Methods:** 700 kidney transplant recipients were prospectively screened for serological responses (median time of 33 (21-52) days) following 3 primary doses of a SARS-CoV2 vaccine. All vaccine doses were received post-transplant, and all 3^rd^ doses were BNT162b2. All participants had serological testing performed post-2^nd^ vaccination at a median time of 34 (IQR 26-46) days following the 2^nd^ inoculation, and at least once prior to their 1^st^ dose of vaccine.

**Results:** 366/700 (52.3%) participants were primed with BNT162b2, whilst 334/700 (47.7) had received ChAdOx1. Overall, 139/700 (19.9%) participants had evidence of prior infection. Of 561 infection naïve participants, 263 (46.9%) had no detectable anti-S following 2-doses of vaccine (V2). 134 (23.9%) participants remained seronegative post 3^rd^ vaccine (V3); 54/291 (18.6%) and 79/270 (29.3%) of participants receiving BNT162b2 and ChAdOx1 respectively, p=0.0029. Median anti-S concentrations were significantly higher post-V3 in patients who had received BNT162b2 compared with ChAdOx1, at 612 (27-234) versus 122 (7.1-1111) BAU/ml respectively, p<0.0001.

Cellular responses were investigated in 30 infection naïve participants at a median time of 35 (24-46) days post-V3. Eighteen of 30 (60.0%) participants had undetectable T-cell responses. There were neither qualitative or quantitative differences in T-cell responses between those patients who received BNT162b2 or ChAdOx1 as their first 2-doses, with 10/16 (62.5%) and 8/14 (57.1%) respectively having undetectable T-cell responses, p=0.77.

**Conclusion:** A significant proportion of transplant recipients remain seronegative following 3 doses of SARS-CoV-2 vaccines, with anti-S concentrations lower in patients receiving heterologous versus homologous vaccinations.

---

Weakened immunogenicity to SARS-CoV-2 vaccines has now been extensively demonstrated in recipients of solid organ transplants(1, 2). To circumvent the potential attenuation in clinical efficacy, many countries have sanctioned additional vaccine doses to transplant and other immunosuppressed populations. Countries leading this strategy are almost exclusively utilising mRNA-based vaccines, either BNT162b2 (Pfizer-BioNTech) or mRNA-1273 (Moderna). Unlike many other developed countries, the UK rolled out BNT162b and ChAdOx1 nCoV-19 replication-deficient adenoviral vector vaccines in equal measure as primary vaccines, with third primary doses, consisting of solely mRNA-based vaccines, approved in September 2021.

Emerging data have since provided evidence on at least equivalent immunogenicity of heterologous compared with homologous vaccine regimens in both transplant recipients and the general population(3-7). In this study, we report on immune responses to 3^rd^ dose BNT162b2 vaccines in transplant recipients either primed with ChAdOx1 or BNT162b as their first two doses.

## Results

Seven-hundred kidney transplant recipients were prospectively screened for serological responses following 3 primary doses of a SARS-CoV2 vaccine. All 3^rd^ doses received were BNT162b2, whilst the first 2 doses were either BNT162b2 or ChAdOx1. All vaccine doses were received post-transplant. All participants had serological testing performed post-2^nd^ vaccination at a median time of 34 (IQR 26-46) days following the 2^nd^ inoculation, and at least once prior to their 1^st^ dose of vaccine. Of 700 participants included, 366/700 (52.3%) had received BNT162b2 as the first 2 doses, whilst 334/700 (47.7) had received ChAdOx1 (*Supplemental Information* **Figure S1**). Overall, 139/700 (19.9%) participants had evidence of prior infection, 75 (20.5%) patients who had received BNT162b2 and 64 (19.2%) patients who had received ChAdOx1, p=0.66.

### Serological responses in infection naïve participants

Of 561 infection naïve participants, 263 (46.9%) had no detectable anti-S following 2-doses of vaccine (V2), with 134 (23.9%) remaining seronegative at a median time of 33 (21-52) days post 3^rd^ vaccine (V3) (**Table 1**). The proportion of participants who were seronegative post-V3 following 3 doses of BNT162b2 remained significantly lower than those who had received ChAdOx1 for the first 2 doses at 54/291 (18.6%) and 79/270 (29.3%) respectively, p=0.0029. On multivariable analysis, clinical characteristics associated with detectable anti-S post-V3 included younger age OR 1.02 (1.00-1.04), p=0.041; receiving calcineurin inhibitor monotherapy as maintenance immunosuppression, OR 4.08 (2.82-8.37), p<0.0001; absence of a diagnosis of diabetes, OR 1.99 (1.27-3.14), p=0.0029; 1^st^ vaccine received after the 1^st^ year post-transplant, OR 4.45 (2.21-9.12), p<0.0001 and receiving BNT162b2 for 1^st^ two vaccines, OR 1.59 (1.03-2.46), p=0.037 (*Supplemental Information* **Table S1**).

**Table 1.**
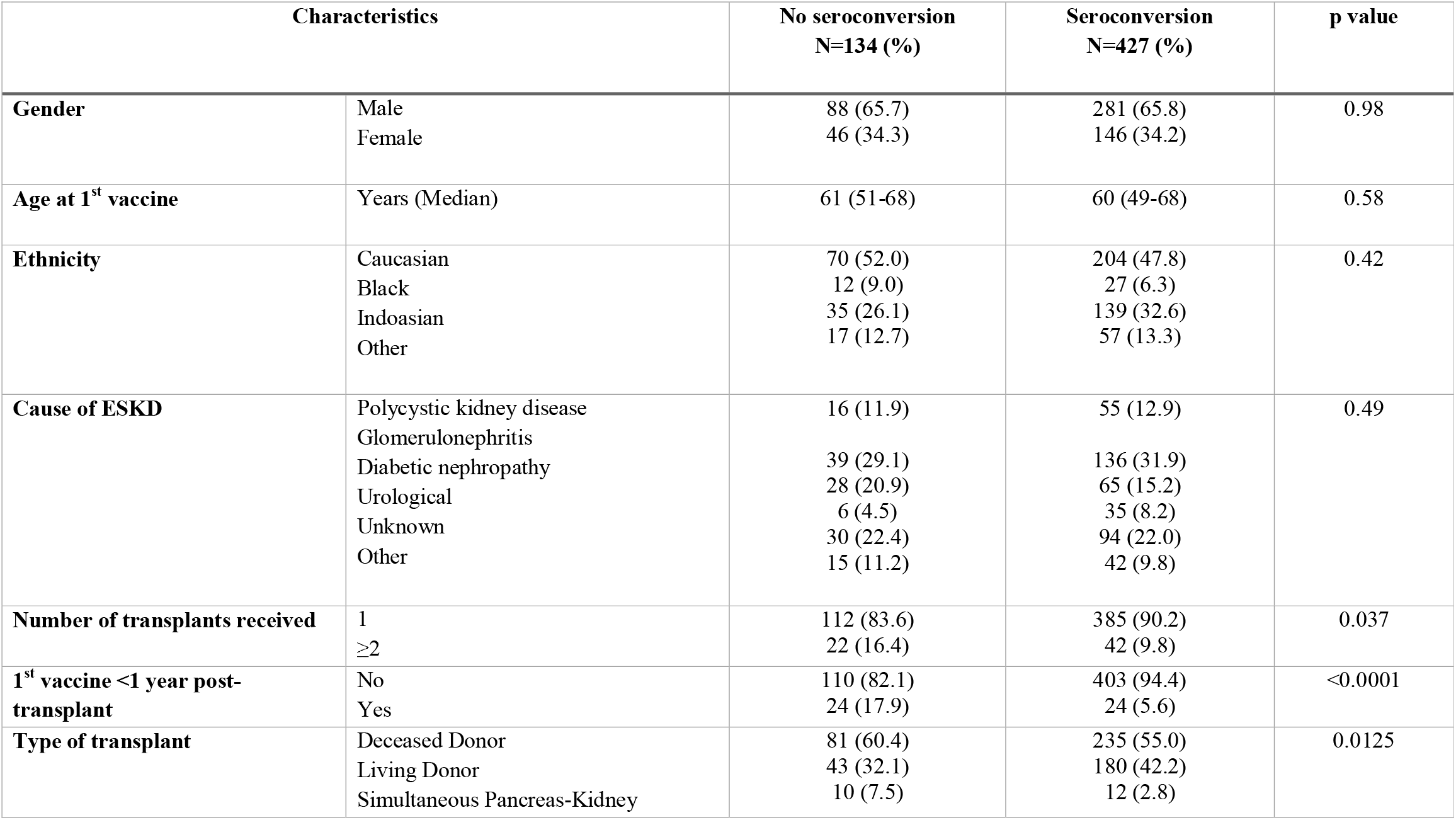
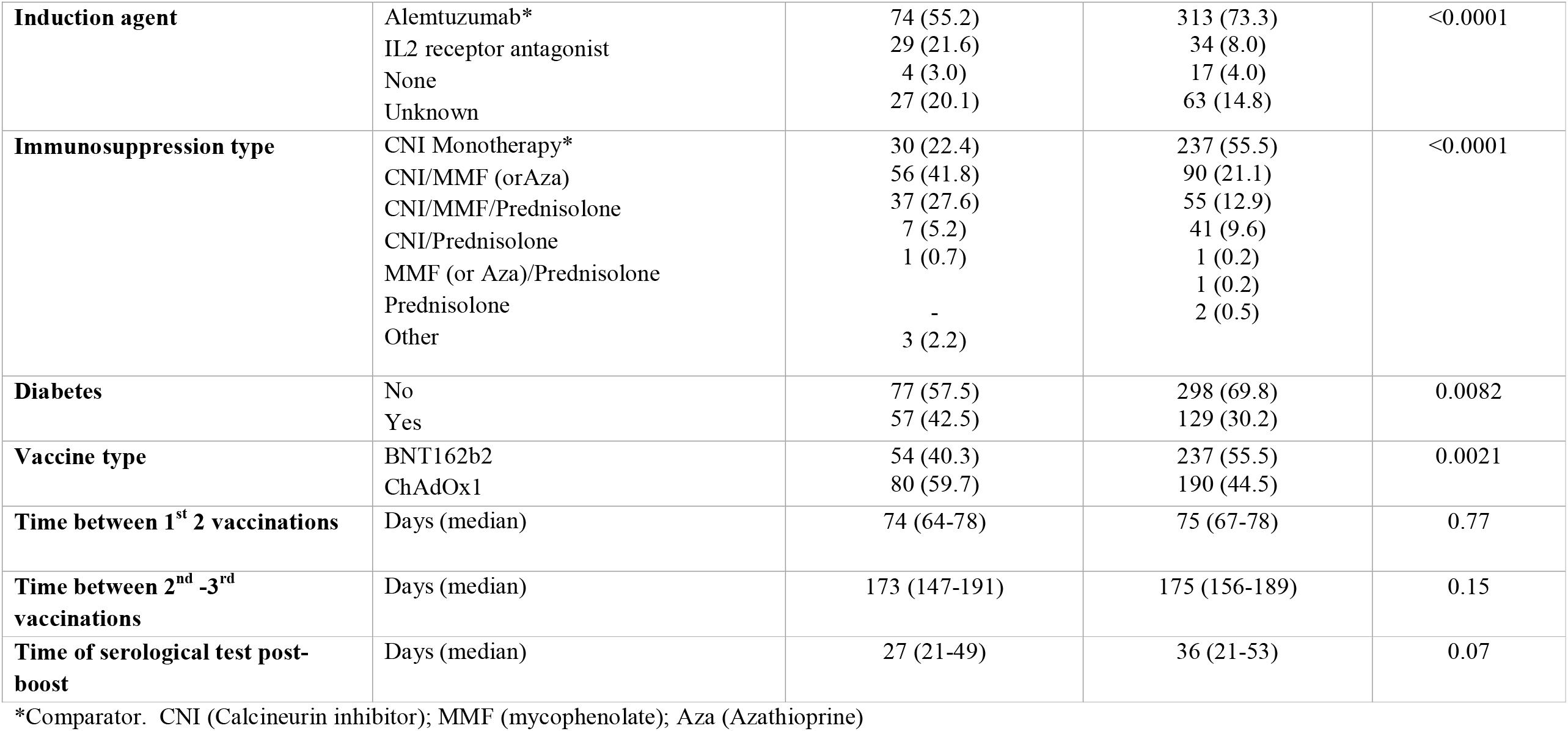
Characteristics between infection-naïve transplant recipients by serostatus following 3^rd^ primary vaccine dose.

Median anti-S concentrations were significantly higher post-V3 in patients who had received BNT162b2 compared with ChAdOx1, at 612 (27-234) versus 122 (7.1-1111) BAU/ml respectively, p<0.0001 (**Figure 1a**). Anti-S concentrations had significantly increased post-V2 compared with V3 in both recipients of BNT162b2, 45 (7.1-510) to 612 (27-234) BAU/ml respectively, p<0.0001, and ChAdOx1, 7.1 (7.1-24) to 122 (7.1-1111) respectively, p<0.0001 (**Figure 1b**).

**Figure 1.**
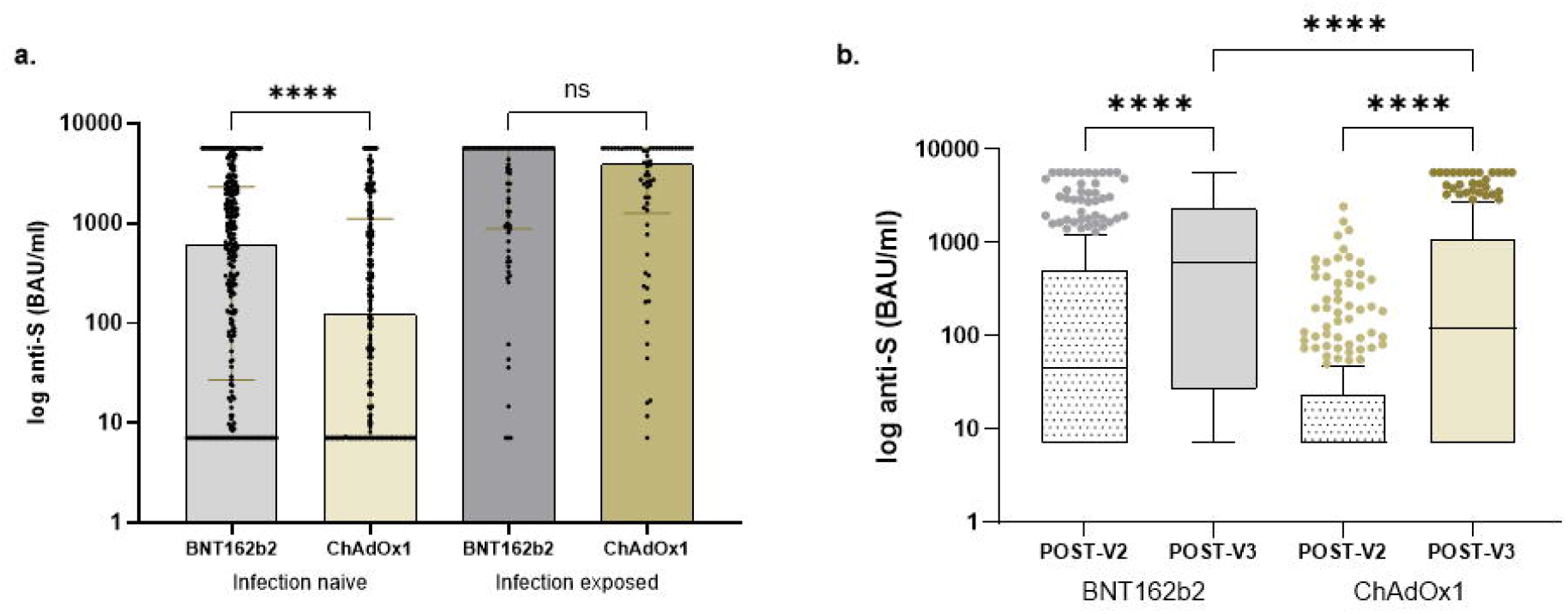
Comparison of anti-S concentrations in transplant recipients. a. Median anti-S concentrations were significantly higher post-V3 in infection-naïve patients who had received BNT162b2 compared with ChAdOx1, at 612 (27-234) versus 122 (7.1-1111) BAU/ml respectively, p<0.0001. There was no difference in anti-S concentrations post-V3 in participants who had received BNT162b2 or ChAdOx1 as their first 2-doses, with concentrations of 5680 (879-5680) compared with 3941 (1255-5680) BAU/ml respectively, p=0.60. b. Anti-S concentrations had significantly increased post-V2 compared with V3 in both recipients of BNT162b2, 45 (7.1-510) to 612 (27-234) BAU/ml respectively, p<0.0001, and ChAdOx1, 7.1 (7.1-24) to 122 (7.1-1111) respectively, p<0.0001

### Serological responses in participants with prior infection

Four of 139 (2.9%) participants defined as having prior exposure remained seronegative post-V3, 3 had RT-PCR proven infection, whilst one participant was anti-NP positive. The median anti-S concentrations in infection exposed participants post-V3 was 4064 (917-5680) BAU/ml). There was no difference in anti-S concentrations post-V3 in participants who had received BNT162b2 or ChAdOx1 as their first 2-doses, with concentrations of 5680 (879-5680) compared with 3941 (1255-5680) BAU/ml respectively, p=0.60 (**Figure 1a**).

### Assessment of cellular responses

Cellular responses were investigated in 30 infection naïve participants at a median time of 35 (24-46) days post-V3. Eighteen of 30 (60.0%) participants had undetectable T-cell responses; clinical characteristics of participants can be found in the *Supplemental Information* (**Table S2**). There was a positive correlation between quantitative serological and cellular responses, r=0.62, p=0.0003. The median anti-S concentrations in ELISpot negative and positive participants were 539 (24-2169) and 2719 (1517-4474) BAU/ml respectively, p=0.0034. There were neither qualitative or quantitative differences in T-cell responses between those patients who received BNT162b2 or ChAdOx1 as their first 2-doses, with 10/16 (62.5%) and 8/14 (57.1%) respectively having undetectable T-cell responses, p=0.77; with median SFU/10^6^ PBMC to S1/S2 peptides of 30 (4-84) and 27 (4-119) respectively, p=0.91. Eleven of the 20 (55.0%) participants who were maintained on CNI monotherapy were ELISpot positive compared with 7/10 (70.0%) of participants on ≥2 immunosuppressive agents, p=0.44; this quantitatively equated to 37 (5-109) and 16 (2-64) SFU/10^6^ PBMC respectively, p=0.29. There was also a trend towards an inverse correlation between increasing age and T-cell response, r=-0.35, p=0.05.

T-cell responses were assessed in 6 participants post-V3 with prior infection, and 5/6 (83.3%) were ELISpot positive. The median SFU/10^6^ PBMC in infection exposed participants was 255 (108-372), which was significantly higher than in the infection naïve group, 28 (4-97) SFU/10^6^ PBMC, p=0.0097.

## Discussion

In this study, we have shown that serological responses to a homologous 3-dose vaccine regimen (BNT162b2 x3) remained superior to a heterologous regimen (ChAdOx1x2/BNT162b x1) in patients without evidence of prior infection, with no differences seen in the measured cellular response between the two groups.

Importantly, a significant number of infection naïve patients, 23.9%, remained seronegative following 3 vaccine doses. Consequently, this population has been offered a 4^th^ or booster vaccine dose in the UK, which was actioned as part of the national response to the Omicron variant. However, to devise ongoing vaccine strategies to optimise the protection of this specific population, there are several aspects to consider. Whilst data suggest incremental increases in seropositivity, and hence anti-S concentrations in transplant recipients, with each consecutive vaccine dose, it is unlikely that patients who remain seronegative after 3-doses will mount a meaningful antibody response after 4-doses(2). This has recently been demonstrated in a 4-dose study of homologous mRNA vaccines, which reported that only 1 in 10 of patients who were seronegative after 3-doses developed an anti-S concentration substantial enough to correlate with the presence of neutralising antibodies, a key predictor of protection against infection. To our knowledge, there are no data on immune responses to 4-doses of vaccine in transplant patients which incorporate a heterologous regimen.

Whilst the majority of studies, including this one, focus predominantly on serological responses, it may be beneficial to consider the T-cell responses in transplant patients, which although less easily measured, may provide a surrogate marker for protection against severe disease(6). This may be of specific relevance to transplant recipients, who are mostly maintained on T-cell directed immunosuppressive agents. In addition, whilst new variants may escape vaccine or naturally acquired antibodies, T-cell responses remain relatively intact, supporting the observation of relative lack of severe disease in vaccinated individuals. It also provides a rationale to target optimisation of cellular responses(8). Although in general, T-cell immunity correlates with B-cell responses, data has shown that cellular immunity may predominate over humoral responses in transplant recipients(3). Consideration of only evidence of serological responses would eliminate the use of viral vector-based vaccines in transplant recipients altogether. This may be premature as evidence suggests that heterologous vaccine doses may result in at least equivocal, if not enhanced, serological and cellular responses in both the general population and transplant recipients(3, 4, 7). Given the relatively small number of patients included in our cellular response analysis, and limitation of assessment of a single functional measure, our conclusions are restricted, but herein we report comparative cellular responses. Therefore, strategies involving multiple doses of heterologous vaccines may still have a role in the mechanism of defence against SARS-CoV-2 infection in transplant recipients.

Whilst immunogenicity data may provide useful data to guide clinical efficacy, real world data on the protection afforded to transplant recipients by vaccination, and the comparative impact of the different vaccines, will be key to the strategic planning to protect transplant recipients. Although data show a reduction in vaccine efficacy across the different platforms in transplant recipients, there are no data to suggest clinical outcomes significantly differ between mRNA-based vaccines and ChAdOx1 after two doses in the era where the Delta variant predominated(10).

Importantly, whilst the transplant community continues to navigate its way through the pandemic, adaptation of approaches to protect transplant recipients will be necessary, and likely to be reliant on a variety of different methods. Such approaches need to be tailored to the individual patients, specific variants, and infection rates. For patients who have inadequate immune responses to 3-doses of vaccine, alternative immune protection could be better offered from passive immunity, however if repeated vaccine dosing is a strategy undertaken for non-responders, further assessment of the merits of heterologous vaccination regimens may be justified.

## Methods

### Study population

The study included 700 kidney transplant recipients, under the care of the Imperial College Renal and Transplant Centre, London. The study ‘The effect of COVID-19 on Renal and Immunosuppressed patients’, sponsored by Imperial College London, was approved by the Health Research Authority, Research Ethics Committee (Reference: 20/WA/0123).

### Serological testing

Serum was tested for antibodies to nucleocapsid protein (anti-NP) using the Abbott Architect SARS-CoV-2 IgG 2 step chemiluminescent immunoassay (CMIA) according to manufacturer’s instructions. This is a non-quantitative assay and samples were interpreted as positive or negative with a threshold index value of 1.4. Spike protein antibodies (anti-S IgG) were detected using the Abbott Architect SARS-CoV-2 IgG Quant II CMIA. Anti-S antibody titres are quantitative with a threshold value for positivity of 7.1 BAU/ml, to a maximum value of 5680 BAU/ml.

### Definition of prior infection

Prior infection was defined serologically or via past PCR positive confirmed infection. The detection of anti-NP on current or historic samples, and the presence of anti-S at baseline (pre-vaccine) or historic samples, was required for the definition of prior infection by serological methods. Prior to December 2021, prior infection was determined by the presence of anti-NP or receptor binding domain (RBD) antibodies, using an in-house double binding antigen ELISA (Imperial Hybrid DABA; Imperial College London, London, UK), which detects total RBD antibodies.

### T cell ELISpot

SARS-CoV-2 specific T-cell responses were detected using the T-SPOT® Discovery SARS-CoV-2 (Oxford Immunotec) according to the manufacturer’s instructions. In brief, peripheral blood mononuclear cells (PBMCs) were isolated from whole blood samples with the addition of T-Cell Select™ (Oxford Immunotec) where indicated. 250,000 PBMCs were plated into individual wells of a T-SPOT® Discovery SARS-CoV-2 plate. The assay measures immune responses to SARS-CoV-2 structural peptide pools; S1 protein, S2 protein, and positive PHA (phytohemagglutinin) and negative controls. Cells were incubated and interferon-γ secreting T cells were detected. Spot forming units (SFU) were detected using an automated plate reader (Autoimmun Diagnostika). Infection-naïve, unvaccinated participants were used to identify a threshold for a positive response using mean +3 standard deviation SFU/10^6^ PBMC, as previously described. This resulted in a cut-off for positivity of 40 SFU/10^6^ PBMC.

### Statistical Analysis

Statistical analysis was conducted using Prism 9.3.1 (GraphPad Software Inc., San Diego, California). Unless otherwise stated, all data are reported as median with interquartile range (IQR). Where appropriate, Mann-Whitney U and Kruskal-Wallis tests were used to assess the difference between 2 or >2 groups, with Dunn’s post-hoc test to compare individual groups. Multivariable analysis was carried out using multiple logistic regression using variables which were found to be significant on univariable analysis.

## Data Availability

All data produced in the present work are contained in the manuscript

## Acknowledgments

This research is supported by the National Institute for Health Research (NIHR) Biomedical Research Centre based at Imperial College Healthcare NHS Trust and Imperial College London. The authors would like to thank the West London Kidney Patient Association, all the patients and staff at ICHNT (The Imperial COVID vaccine group and dialysis staff, and staff within the North West London Pathology laboratories). The authors are also grateful for support from The Nan Diamond Fund, Sidharth and Indira Burman, and the Auchi Charitable Foundation. MP is supported by an NIHR clinical lectureship. Work in DCT’s lab is supported by a Wellcome Trust Clinical Career Development Fellowship

