## Supplemental Information for "Comparative immunogenicity of heterologous versus homologous 3rd SARS-CoV-2 vaccine doses in kidney transplant recipients"

**Figure S1. Study Flow Diagram**

**Inclusion Criteria**

- All post-V3 (with BNT162b2 as 3^rd^ dose)
- All vaccinations occurred post-transplant
- All participants had serological screening post-V2 and at least once pre-1^st^ vaccine

Study Participants n=700

ChAdOx1 n=334 (47.7%)

Prior infection n=64 (19.2%)

Infection naive n=270 (80.8%)

BNT162b2 n=366 (52.3%)

Prior infection n=75 (20.5%)

Infection naive n=291 (79.5%)

**Table S1. Clinical characteristics associated with non-seroconversion following 3^rd^ primary dose vaccinations in infection naïve transplant recipients**

| **Variable** | **Reference Group** | **Univariable** | | **Multivariable** | |
| --- | --- | --- | --- | --- | --- |
|  |  | **OR (95% CI)** | **P value** | **OR (95% CI)** | **P value** |
| Age |  | 0.99 (0.98-1.00) | 0.0007 | 0.98 (0.96-0.99) | 0.041 |
| Number of transplants | 1 | 0.56 (0.32-0.98) | <0.0001 | 1.24 (0.66-2.29) | 0.49 |
| Type of graft | Deceased Donor | 1.54 (1.03-2.34) | 0.039 | 1.38 (0.87-2.21) | 0.17 |
| Induction agent | Alemtuzumab | 0.45 (0.30-0.67) | <0.0001 | 0.85 (0.53-1.38) | 0.51 |
| Immunosuppression | CNI monotherapy | 0.23 (0.15-0.36) | <0.0001 | 0.21 (0.12-0.35) | <0.0001 |
| Diabetes | No | 0.58 (0.39-0.87) | <0.0001 | 0.50 (0.32-0.79) | 0.0029 |
| Vaccine | BNT162b2 | 0.54 (0.36-0.80) | 0.0023 | 0.63 (0.41-0.97) | 0.037 |
| Vaccine within the 1^st^ year post-transplant | No | 0.27 (0.15-0.50) | <0.0001 | 0.22 (0.11-0.45) | <0.0001 |

**Table S2. Clinical characteristics of infection-naïve participants by T-cell responses post-V3**

| Characteristics | | ELISpot negative  N=18 (%) | ELISpot positive  N=12 (%) | p value |
| --- | --- | --- | --- | --- |
| Gender | Male  Female | 14 (77.8)  4 (22.2) | 10 (83.3)  2 (16.7) | 0.71 |
| Age at 1^st^ vaccine | Years (Median) | 62 (60-75) | 55 (46-64) | 0.07 |
| Ethnicity | Caucasian  Black  Indoasian | 9 (50.0)  2 (11.1)  7 (38.7) | 4 (33.3)  1 (8.3)  7 (58.3) | 0.58  0.37 |
| Cause of ESKD | Polycystic kidney disease  Glomerulonephritis  Diabetic nephropathy  Urological  Unknown  Other | 3 (16.7)  6 (33.3)  1 (5.6)  1 (5.6)  5 (27.8)  2 (11.1) | 1 (8.3)  4 (33.3)  4 (33.3)  -  3 (25.0)  - | 0.33 |
| Number of transplants received | 1  ≥2 | 16 (88.9)  2 (11.1) | 12 (100) | 0.24 |
| 1^st^ vaccine <1 year post-transplant | No  Yes | 17 (94.4)  1 (5.6) | 12 (100) | 0.41 |
| Type of transplant | Deceased Donor  Living Donor | 16 (88.9)  2 (11.1) | 5 (41.7)  7 (58.3) | 0.007 |
| Induction agent | Alemtuzumab  IL2  Unknown | 14 (77.8)  2 (11.1)  2 (11.1) | 11 (91.7)  -  1 (8.3) | 0.46 |
| Immunosuppression type | CNI Monotherapy*  CNI/MMF (orAza)  CNI/MMF/Prednisolone  CNI/Prednisolone | 11 (61.1)  3 (16.7)  1 (5.6)  3 (16.7) | 9 (75.0)  -  -  3 (25.0) | 0.37 |
| Diabetes | No  Yes | 15 (83.3)  3 (16.7) | 6 (50.0)  6 (50.0) | 0.06 |
| Vaccine type | BNT162b2  ChAdOx1 | 10 (55.6)  8 (44.4) | 6 (50.0)  6 (50.0) | 0.77 |
| Anti-S | Positive  Negative | 16 (88.9)  2 (11.1) | 12 (100) | 0.24 |
| Anti-S concentrations | Median (BAU/ml) | 539 (24-2169) | 2719 (1517 -4474) | 0.0034 |
